# Progression and trajectory network of age-related functional impairments and their associations with mortality: a two-decade prospective study

**DOI:** 10.1101/2022.08.25.22279239

**Authors:** Hui Chen, Binghan Wang, Rongxia Lv, Tianjing Zhou, Jie Shen, Huan Song, Xiaolin Xu, Yuan Ma, Changzheng Yuan

**Author notes:** **Corresponding author:** Changzheng Yuan, School of Public Health, Zhejiang University School of Medicine, Hangzhou, Zhejiang, 310058. HC and BW had equal contributions as co-first authors.

## Abstract

**Objective:** To characterize the progression and trajectory network of age-related functional impairments and assess their associations with mortality.

**Design:** Prospective cohort study.

**Setting:** The Health and Retirement Study (HRS, 2000-2020).

**Participants:** A total of 17 914 HRS participants aged from 51 to 90 years in 2000.

**Main outcome measure:** Age-related functional impairments including visual impairment, hearing impairment, cognitive impairment, physical frailty, restless sleep, and depression, all measured using structural questionnaires biennially or quadrennially. All-cause mortality was ascertained from multiple sources.

**Results:** During follow-up (median=12 years), the incidence rates of visual impairment (59.3 cases/1000 person years), hearing impairment (52.1 cases/1000 person years), physical frailty (31.7 cases/1000 person years), and cognitive impairment (42.5 cases/1000 person years) increased exponentially with age (*P*-trend <0.001), while those of restless sleep (75.6 cases/1000 person years) and depression (35.6 cases/1000 person years) increased relatively slowly. We detected bidirectional associations among all ARFIs (*P* <0.001). Overall, each additional ARFI was associated with 13% (11%-15%) higher risk of mortality, with independent associations observed for physical frailty (hazard ratio: 1.59, 95% confidence interval: 1.49-1.69), depression (1.38, 1.30-1.46), visual impairment (1.19, 1.13-1.26), and cognitive impairment (1.13, 1.06-1.21).

**Conclusions:** ARFIs were highly interconnected as a network and were associated with mortality, which highlighted the importance of integrated strategies to monitor and manage the ARFIs to achieve healthy longevity.

**SUMMARY BOXES:** *WHAT IS ALREADY KNOWN ON THIS TOPIC:*

- Aging is characterized by changes in multiple functions, such as visual, hearing, and cognitive impairments.
- Specific age-related functional impairments are associated bidirectionally.

*WHAT THIS STUDY ADDS:*

- In a prospective study, the incidence rates of visual impairment, hearing impairment, physical frailty, and cognitive impairment increased exponentially as age increased, while incidence rates of restless sleep and depression increased relatively slowly with age.
- The six ARFIs are bidirectionally related to each other and predicted higher risk of mortality in a dose-response manner, with independent associations observed for visual impairment, cognitive impairment, physical frailty, and depression.

## INTRODUCTION

In the context of worldwide population aging,^1 2 3^ healthy longevity has become increasingly critical.^4 5^ Aging is characterized by changes in multiple functions, including sensory, physical, mental, and cognitive function,^6 7^ which largely influence the independence, life quality, and longevity of older adults. Age-related functional impairments (ARFIs) often co-exist and their incidence and progression are highly intertwined.^8 9 10 11^ For example, physical frailty was associated with a higher risk of cognitive impairment, disability, and death,^7 12 13^ and cognitive decline was related to an increased risk of falling and frail symptoms.^14^ To further understand the aging process and identify effective preventive measures of ARFIs, there warrant investigations into their complex and interactive relations.

Until now, studies have made exploratory efforts in constructing the trajectory networks of aging-related diseases^15 16 17 18^ in the field of multimorbidity. For instance, a previous study in the Danish National Patient Register presented a trajectory network in a single connected graph to display the multimorbidity spectrum of a specific disease.^16^ Another study using follow-up data from a clinical trial examined the state transitions among age-related diseases using multistate models.^19^ However, these studies all focused on disease diagnosis, while there are few attempts to uncover the trajectory networks of first occurrences of ARFIs, which often requires frequent and long-term follow-ups and comprehensive assessments of the functions of interest. Also, although previous studies have identified the associations of some ARFIs with mortality,^13 20 21 22 23 24 25^ whether they are independently related to mortality and their combined relations remained largely unknown.

Therefore, we aimed to characterize the trajectory networks of age-related functional impairments and their associations with mortality in the Health and Retirement Study (HRS), a nationally representative prospective study with two decades of biennial follow-ups.

## METHODS

### Study population

The HRS is a national longitudinal study among middle-aged or older adults in the United States. Participants born in 1931-1941 were recruited in 1992 (HRS original cohort). In 1993/1994, participants born before 1924 were included in the Asset and Health Dynamics Among the Oldest Old (AHEAD) cohort. In 1998, participants born in 1924-1930 (Children of the Depression, CODA) and 1942-1947 (War Babies, WB) were enrolled. The four non-overlapping cohorts formed a nationally representative sample of US adults aged 50 years or older. We stratified our analysis by sub-cohorts where applicable to represent age differences. Participants were revisited biennially through telephone, Internet, or face-to-face interviews. The overall response rate was approximately 85%.^26^ More detailed information can be found elsewhere (https://hrs.isr.umich.edu/). This HRS was approved by the Institute for Social Research of University of Michigan (IRB Protocol: HUM00061128). Written informed consent was obtained from all participants or proxy.

In the current study, we included 17 914 participants aged from 51 to 90 years with available data of ARFIs in 2000 and followed them up until 2020. We conducted several analyses with different sample sizes (**Supplemental Figure S1**): (1) we included participants without corresponding ARFIs at baseline to calculate their incidence rates (n=12 756 participants for visual impairment, 12 974 for hearing impairment, 11 028 for cognitive impairment, 14 346 for physical frailty, 9671 for restless sleep, and 12 786 for depression); (2) we included 15 852 participants without any missing ARFI measurement at baseline to describe the progression of co-existing ARFIs; (3) we included participants without missing values of ARFIs or the corresponding outcome ARFIs at baseline to construct the hazard trajectory network (n=13 913 when treating visual impairment as the exposure, 14 270 for hearing impairment, 13 609 for cognitive impairment, 15 874 for physical frailty, 10 833 for restless sleep, and 14 247 for depression).

### Standard Protocol Approvals, Registrations, and Patient Consents

No patients were involved in setting the research question or the outcome measures, nor were they involved in the design and implementation of the study.

### Assessments of age-related functional impairments and ascertainment of mortality

In this study, ARFIs included visual impairment, hearing impairment, restless sleep, cognitive impairment, physical frailty, and depression, all measured biennially except for physical frailty (quadrennially). Participants who rated their visual or hearing functions as fair or poor were defined as having visual or hearing impairment.^27^ Similarly, participants were asked whether their sleep was restless (yes or no). According to a previous study in the HRS which modified the Fried criteria, participants aged >=65 years were physically frail if they met more than one of the Fried criteria: low level of physical activity, exhaustion, slowness, weakness, and weight loss.^28 29^ Cognitive function was measured using tests adapted from the Telephone Interview for Cognitive Status (TICS) in a 27-point scale,^30^ including an immediate and a delayed 10-noun free recall test (1 point for each), serial 7 subtractions (i.e., subtract 7 from 100 and continue subtracting 7 from each subsequent number for a total of five trials, 1 point for each trial), and backward counting from 20 (2 points). A global cognitive score lower than 12 indicated cognitive impairment.^30^ The validated 8-item Center for Epidemiologic Studies-Depression (CES-D) scale was used to assess depression.^31^ Considering the shared component of CES-D and sleep quality, we excluded the sleep item from the scale and a total score >=4/7 indicated depression.^32 33^ All-cause mortality was ascertained using data linkage from the population registry and through interview with informants or knowledgeable others

### Other Covariates

We included multiple covariates for confounder adjustments. Sociodemographic factors included age, gender defined by self-identity, race (White / Caucasian / Black / Others), body weight (kg), height (m), and education level (lower than high school / general educational development (GED) / high school / college / above). Lifestyle factors included smoking status (never / former / current), alcohol consumption (never / former / current), household income, and vigorous exercise (whether more than three times a week). Health conditions included stroke, cancer, memory-related diseases, and other psychological diseases.

### Statistical Analyses

Baseline characteristics of the study population were described by birth cohorts (WB, HRS original cohort, CODA, and AHEAD). Continuous variables were presented in mean (standard deviation) and categorical variables were presented in number (percentage). Because physical frailty was measured every four years, we calculated the four-year incidence rates (2000-2004) of the six ARFIs by age and sex. We characterized their co-existing patterns by birth cohorts at baseline using the ‘UpSetR’ package.^34^ We used the Sankey diagram to characterize the status transitions of participants from 2000 to 2020 using the ‘ggalluvial’ package.^35^

To illustrate how the ARFIs are associated with each other, we constructed a hazard trajectory network following a three-step strategy introduced in a previous study.^22^ First, we calculated the hazard ratios (HRs) and 95% confidence intervals (CIs) for the directional association of each ARFI with other ARFIs and all-cause mortality using Cox proportional hazard models. The person-time was calculated from baseline to the first occurrence of outcome of interest, death, or loss-to-follow-up, whichever came first. Models were adjusted for age, gender, BMI, race, education, family income, smoking status, drinking status and frequency of vigorous exercise at baseline. The associations reaching statistical significance at the Bonferroni-corrected thresholds were carried to the second step, where the estimates were further adjusted for prevalence and incidence of other ARFIs to eliminate indirect associations (i.e., that were confounded or mediated by other ARFIs). The associations remaining statistically significant were considered independent links between ARFIs. Using a directed network graph, we integrated these independent links to form a hazard trajectory network. For example, when assessing the association of visual impairment with cognitive impairment, we first calculated the multivariable-adjusted HR for incident cognitive impairment associated with baseline visual impairment. If the association was statistically significant, we would further adjust the model for prevalence and incidence of other ARFIs (in this case, hearing impairment, physical frailty, restless sleep, and depression, categorized as prevalent, incident or neither) to test whether the relation of visual impairment to cognitive impairment was independent of other ARFIs. Finally, if the association of visual impairment with cognitive impairment was independent of other ARFIs, it would be shown on the trajectory network as an arrow from visual impairment to cognitive impairment. We further assessed the association of the number of ARFIs with mortality using Cox proportional hazard models.

We performed a series of sensitivity analyses. First, we excluded people with prevalent stroke, cancer, memory-related diseases, or other psychological diseases that might be directly linked to functional changes and mortality. We excluded such patients only in the sensitivity analysis because our intention was to depict the progression of ARFIs in the general population rather than only healthy participants. To further assess the trajectory network in relatively healthier participants, we restricted our analyses to participants with none of the six ARFIs at baseline. Third, considering that participants who were at risk of developing ARFIs were also at higher risk of mortality, we treated all-cause death as the competing risk using the Fine and Gray model.^36^

All statistical analyses were completed using R 4.2.0. All *P*-values were two-sided and Bonferroni-corrected *P* < 0.05 was considered an indicator of statistical significance to address multiplicity.

## RESULTS

### Participants Characteristics

Among the 17 914 participants included, 57.5% were female, 82.9% were White/Caucasian, and the mean (SD) baseline age was 67.5 (9.6) years (**Table 1**). Participants who were older were more likely to be female, less educated, never alcohol drinkers, never smokers, and less physically active. For example, compared to the WB participants (mean [SD] age 54.8 [2.8] years old), AHEAD participants (mean [SD] age 80.2 [5.2] years old) were more likely to be less educated (1651 [36.6%] *vs* 290 [14.4%] with lower than high school education), non-drinkers (2959 [65.6%] *vs* 884 [43.9%]), never smokers (2123 [47.3%] *vs* 791 [39.3%]), and less physically active (1378 [30.9%] *vs* 991 [49.2%]).

**Table 1.** Baseline Characteristics of the Study Participants in the Health and Retirement Study

| Characteristics | Overall<br>(N = 17 914) | Birth cohorts |  |  |  |
| --- | --- | --- | --- | --- | --- |
|  |  | WB<br>(n = 2015) | HRS<br>(n = 9315) | CODA<br>(n = 2071) | AHEAD<br>(n = 4513) |
| Age, years (mean (SD)) | 67.5 (9.6) | 54.8 (2.8) | 63.0 (4.9) | 72.3 (2.2) | 80.2 (5.2) |
| Female, N (%) | 10 300 (57.5) | 975 (48.4) | 5083 (54.6) | 1249 (60.3) | 2993 (66.3) |
| Race, N (%) |  |  |  |  |  |
| White race | 14 852 (82.9) | 1618 (80.3) | 7564 (81.2) | 1797 (86.8) | 3873 (85.8) |
| Black race | 2442 (13.6) | 303 (15.0) | 1405 (15.1) | 185 (8.9) | 549 (12.2) |
| Others | 615 (3.4) | 93 (4.6) | 343 (3.7) | 89 (4.3) | 90 (2.0) |
| Education, N (%) |  |  |  |  |  |
| Lower than High-School | 4872 (27.2) | 290 (14.4) | 2339 (25.1) | 592 (28.6) | 1651 (36.6) |
| GED | 770 (4.3) | 100 (5.0) | 467 (5.0) | 81 (3.9) | 122 (2.7) |
| High school | 5666 (31.6) | 584 (29.0) | 3003 (32.2) | 646 (31.2) | 1433 (31.8) |
| Some college | 3433 (19.2) | 487 (24.2) | 1808 (19.4) | 384 (18.5) | 754 (16.7) |
| College and above | 3173 (17.7) | 554 (27.5) | 1698 (18.2) | 368 (17.8) | 553 (12.3) |
| BMI, kg/m<sup>2</sup> (mean (SD)) | 27.1 (5.3) | 28.4 (5.8) | 27.7 (5.4) | 26.8 (4.9) | 25.25 (4.7) |
| Income, dollars (mean (SD)) | 14 741.9 (59 065.1) | 21 637.9 (81 384.0) | 17 374.1 (69 236.2) | 9 459.6 (25 650.5) | 8 654.2 (24 723.7) |
| Drinking status, N (%) |  |  |  |  |  |
| Never | 9826 (54.9) | 884 (43.9) | 4817 (51.7) | 1166 (56.3) | 2959 (65.6) |
| Former | 3257 (18.2) | 442 (21.9) | 1735 (18.6) | 385 (18.6) | 695 (15.4) |
| Current | 4827 (27.0) | 689 (34.2) | 2761 (29.6) | 520 (25.1) | 857 (19.0) |
| Smoking status, N (%) |  |  |  |  |  |
| Never | 7211 (40.6) | 791 (39.3) | 3437 (37.4) | 860 (41.5) | 2123 (47.3) |
| Former | 7882 (44.4) | 777 (38.6) | 4063 (44.2) | 972 (46.9) | 2070 (46.2) |
| Current | 2667 (15.0) | 447 (22.2) | 1690 (18.4) | 239 (11.5) | 291 (6.5) |
| Vigorous physical activity, N (%) <sup>a</sup> | 7570 (42.3) | 991 (49.2) | 4360 (46.8) | 878 (42.4) | 1341 (29.7) |
| Visual impairment, N (%) | 3900 (21.9) | 332 (16.5) | 1776 (19.1) | 414 (20.1) | 1378 (30.9) |
| Hearing impairment, N (%) | 3629 (20.3) | 243 (12.1) | 1585 (17.0) | 437 (21.1) | 1364 (30.3) |
| Cognitive impairment, N (%) | 4589 (27.8) | 285 (15.1) | 1692 (19.7) | 668 (34.2) | 1944 (47.4) |
| Physical frailty, N (%) | 2026 (11.3) | 115 (5.7) | 840 (9.0) | 243 (11.7) | 828 (18.4) |
| Restless sleep, N (%) | 5362 (33.1) | 634 (33.9) | 2788 (32.7) | 621 (32.3) | 1319 (33.9) |
| Depression, N (%) | 1948 (12.0) | 214 (11.5) | 955 (11.2) | 209 (10.9) | 570 (14.6) |

BMI: body mass index, WB: War Baby cohort born in 1942-1947, HRS: HRS original cohort born in 1931-1941, CODA: Children of Depression cohort born in 1924-1930, AHEAD: Asset and Health Dynamics Among the Oldest Old cohort born before 1924.
<sup>a</sup> Vigorous physical activity includes running, jogging, swimming, cycling, aerobics or gym workout, tennis, digging, and it had been defined as “yes” when it was more often than three times a week.

### Overall, age- and sex-specific four-year incidence rates of ARFIs

During the study follow-up (median [interquartile range] 12 [6-18] years) of all participants, 5934 incident visual impairment cases, 5306 incident hearing cases, 4951 incident restless cases, 6656 incident cognitive impairment cases, 5431 incident physical frailty, and 3679 incident depression cases were identified. The four-year incidence rates were 59.3 cases/1000 person-years for visual impairment, 52.1 for hearing impairment, 42.5 for cognitive impairment, 31.7 for physical frailty, 75.6 for restless sleep, and 35.6 for depression (**Supplemental Table S1**).

The incidence rates of visual impairment, hearing impairment, cognitive impairment, and physical frailty were exponentially higher in older ages (*P*-trends < 0.001 and *P*-nonlinear <0.05; **Figure 1A**), while those for depression and restless sleep increased relatively slowly. For example, the incidence rate for cognitive impairment was 45.0 cases/1000 person-years for participants aged 66-70 years old and 91.5 cases/1000 person-years for participants aged 76-80 years old. The incidence rate of restless sleep was the highest among all ARFIs before the age of 75 years but was surpassed by cognitive impairment in older age groups.

**Figure 1.**
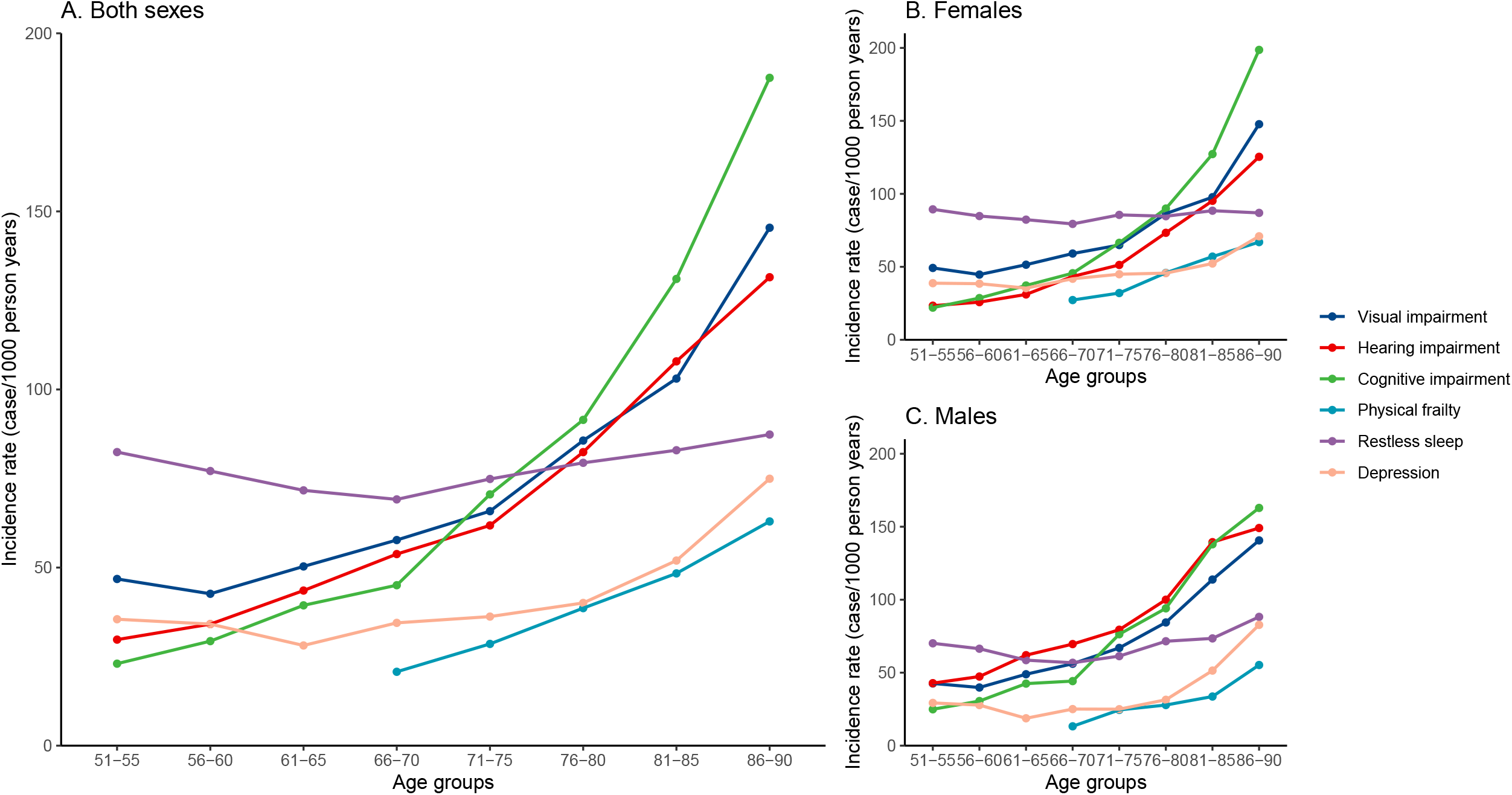
Age-specific incident rates of age-related functional impairments, in overall participants (A), females (B) and males (C) Physical frailty indicators are only measured among participants aged over 65.

We observed similar trends for women and men (**Figure 1B, C**). Female participants had higher incidence rates of restless sleep, physical frailty, and depression (84.3 *vs* 63.6 cases/1000 person-years for restless sleep, 38.4 *vs* 22.8 cases/1000 person-years for physical frailty, 41.4 *vs* 29.3 cases/1000 person-years for depression, *P* < 0.001 for all), while male participants had higher incidence rate of hearing impairment (67.4 *vs* 43.1 cases/1000 person-years, *P* < 0.001).

### Co-existence and progression of ARFIs

A total of 4529 (28.6%) participants had co-existing ARFIs in 2000, and the proportions were 20.1% in WB, 24.0% in HRS, 30.2% in CODA, and 42.0% in AHEAD. Restless sleep and visual impairment were the most prevalent co-existing pair (1469, 9.3%), followed by restless sleep and depression (1307, 8.2%), restless sleep and hearing impairment (1179, 7.4%), and visual and hearing impairment (1116, 7.0%). Patterns of co-existing ARFIs also change with age (**Supplemental Table S2, Figure S2**). For example, in the youngest cohort (WB), the pair of visual impairment restless sleep (8.5%) was the most prevalent, while in the oldest cohort (AHEAD), the visual and hearing impairment pair was the most prevalent (11.9%).

From 2000 to 2020, an increasing proportion of surviving respondents had co-existing ARFIs (from 28.6% to 64.2%; **Supplemental Table S3**). Similar trends were observed for all birth cohorts (**Figure 2**). For example, the proportion of WB participants with co-existing ARFIs increased from 20.1% (367) to 55.1% (374), and that for the AHEAD cohort increased from 42.0% (1582) to 90.5% (114).

**Figure 2.**
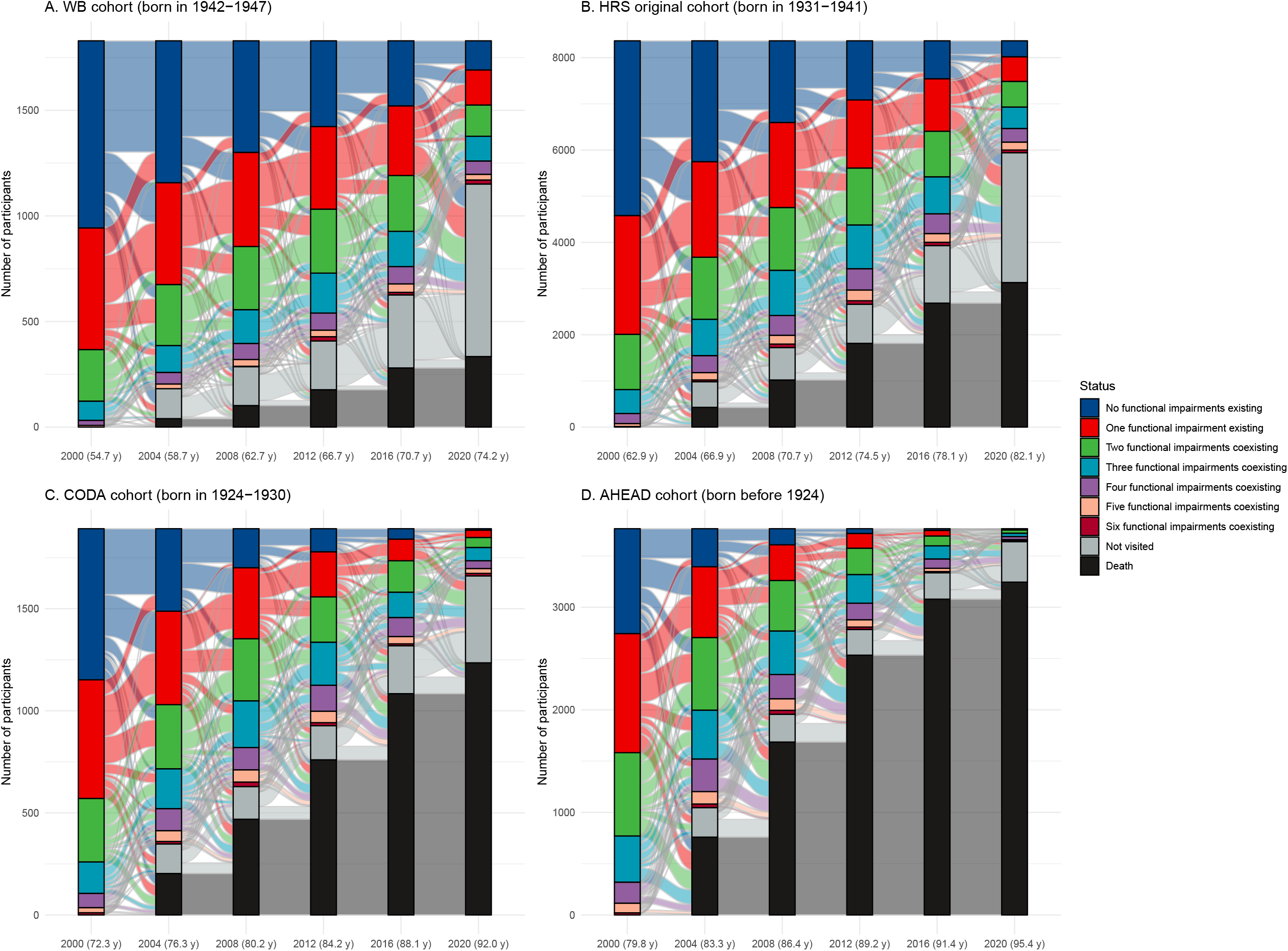
Progression of co-existence of age-related functional impairments from 2000 to 2020 by birth cohorts. WB cohort: War Baby cohort born 1942 to 1947; HRS cohort: HRS original cohort born 1931 to 1941; CODA cohort: Children of Depression cohort born 1924 to 1930; AHEAD cohort: Asset and Health Dynamics Among the Oldest Old cohort born before 1924. Labels of the x-axis were expressed as the year (mean age).

### Hazard trajectory network of functional impairments

In the hazard trajectory network presented in **Figure 3**, all six ARFIs were significantly related to each other. For example, visual impairment predicted higher risks of hearing impairment (HR 2.48, 95% CI 2.29-2.69), depression (2.18, 2.00-2.39), physical frailty (2.08, 1.92-2.24), restless sleep (1.72, 1.59-1.86), and cognitive impairment (1.63, 1.53-1.74), and cognitive impairment showed relations to visual impairment (1.72, 1.57-1.88), hearing impairment (1.43, 1.30-1.57), depression (1.50, 1.35-1.68), physical frailty (1.56, 1.42-1.71), and restless sleep (1.25, 1.13-1.38). More details are shown in **Supplemental Table S4**.

**Figure 3.**
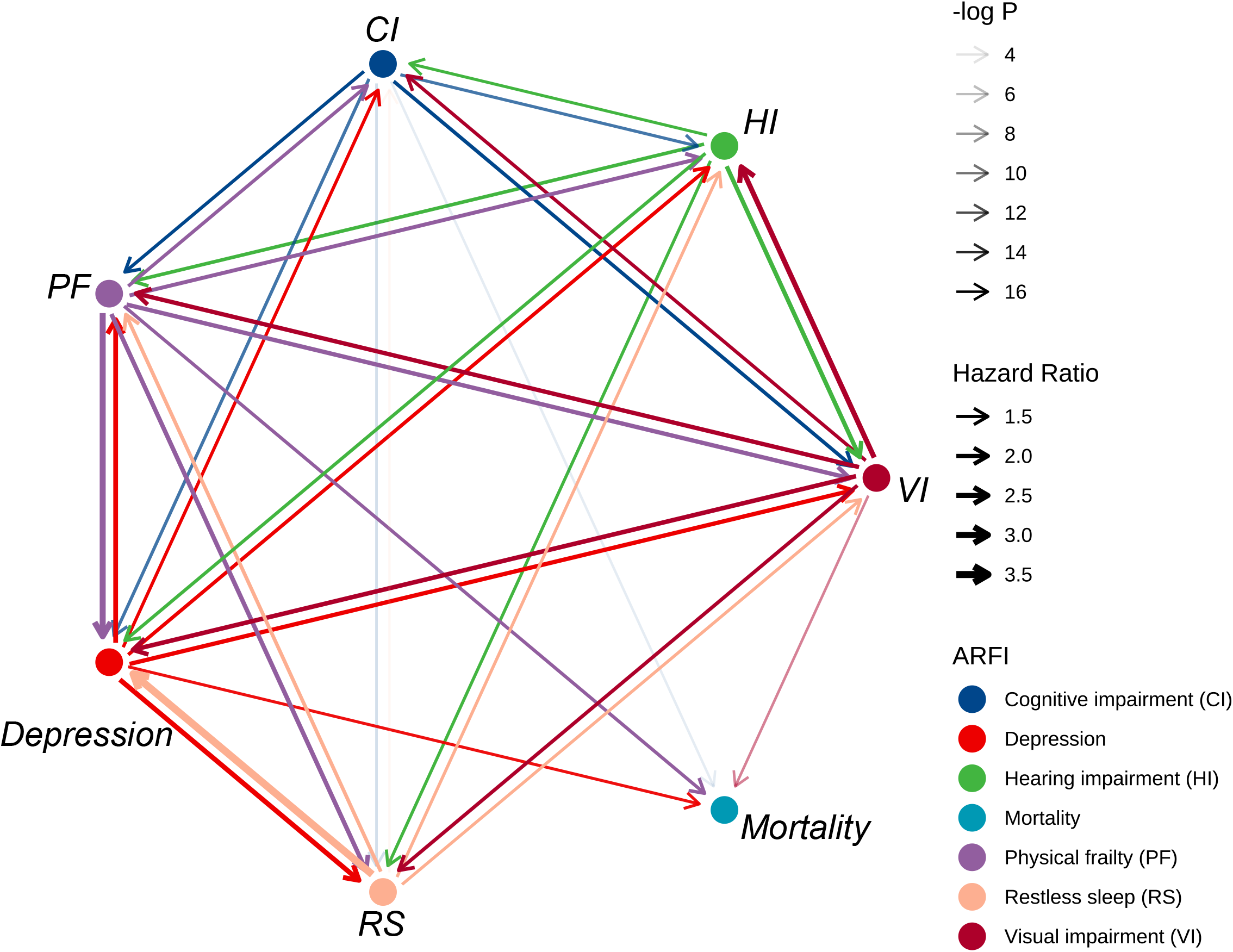
Hazard trajectory networks of age-related functional impairments in the Health and Retirement Study. ARFI: age-related functional impairment, VI: visual impairment, HI: hearing impairment, CI: cognitive impairment, PF: physical frailty, RS: restless sleep. The directed edges indicated the directed associations between ARFIs (nodes), the transparency represented the statistical significance (the more transparent, the less significant), and the thickness indicated the magnitude of relations (the thicker arrow represented larger HR. All analyses were adjusted for age, gender (male / female), BMI, race (White/Caucasian / Black / Others), education (lower than high school / general educational development (GED) / high school / college / above), family income, smoking status (never / former / current), drinking status (never / former / current) and vigorous exercise (whether more than three times a week) at baseline and other baseline and incident ARFIs.

Moreover, a higher number of ARFIs was related to higher risk of mortality, with the HR (95% CI) for each additional ARFI being 1.13 (1.11-1.15). Compared to participants with no ARFI, participants with one, two, three, and more ARFIs were at 12% (6%-18%), 35% (26%-44%), 55% (43%-68%), 43% (27%-61%), and 75%, (50%-104%) higher mortality risk, respectively (**Figure 4**). Furthermore, physical frailty, depression, visual impairment, and cognitive impairment each independently predicted 59% (49%-69%), 38% (30%-46%), 19% (13%-26%) and 13% (6%-21%) higher risk of all-cause mortality.

**Figure 4.**
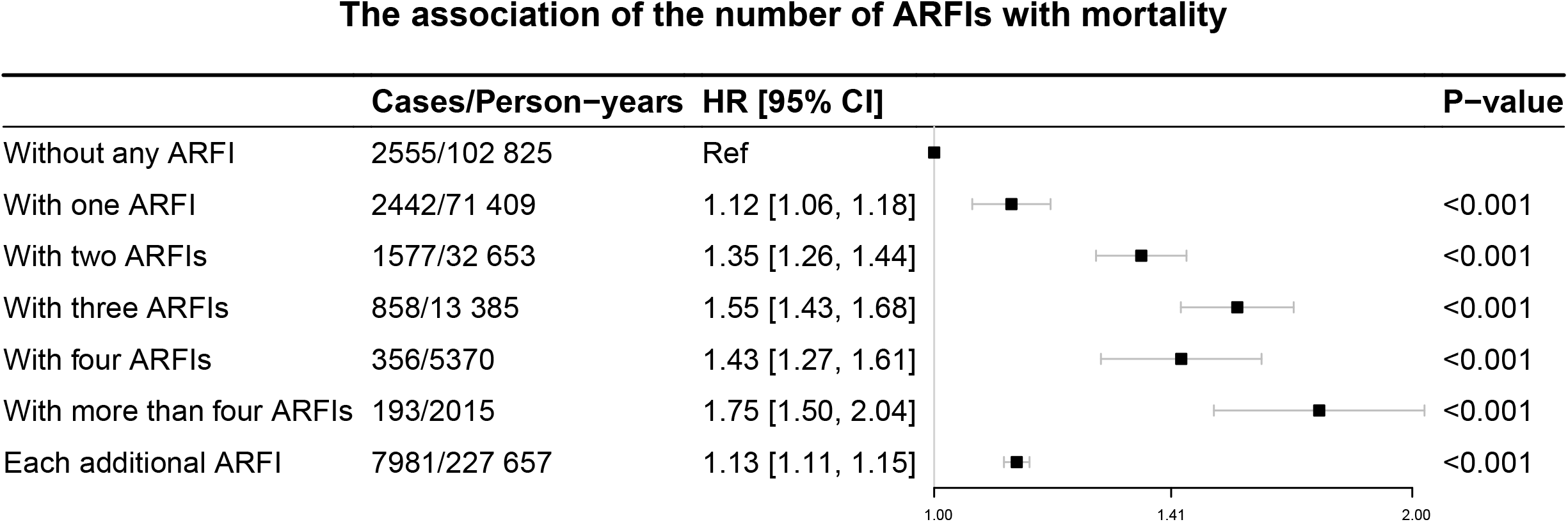
Hazard ratios (HRs) and 95% confidence intervals (CIs) of mortality according to the number of age-related functional impairment (ARFIs) in the Health and Retirement Study. The HRs were adjusted for age, gender (male / female), BMI, race (White/Caucasian / Black / Others), education (lower than high school / general educational development (GED) / high school / college / above), family income, smoking status (never / former / current), drinking status (never / former / current) and vigorous exercise (whether more than three times a week) at baseline.

In the sensitivity analyses (**Supplemental Table S5, Figure S3**), the exclusion of participants with baseline stroke, cancer, memory-related diseases, and other psychological diseases did not substantially alter the trajectory network. When we restricted our analyses to 6373 participants without any ARFIs at baseline, the directions of the associations remained similar and significant. When treating mortality as a competing risk, the trajectory network remained generally similar as in the primary analysis.

## DISCUSSION

In this prospective study with two decades of follow-up, we showed that age-related functional impairments are highly intertwined as a whole. The incidence rates of visual impairment, hearing impairment, cognitive impairment, and physical frailty increased exponentially with age. As age increased, the prevalence and complexity of co-existing ARFIs also increased. We constructed a hazard trajectory network, which showed that the ARFIs were all bidirectionally associated. A larger number of ARFIs was related to higher all-cause mortality risk in a dose-response manner, with independent associations observed for visual impairment, physical frailty, cognitive impairment, and depression.

The incidence rates reported in this study were comparable with previous studies.^37 38 39 40 41 42 43 44 45^ For instance, the incidence rate of cognitive impairment was 52.6 cases/1000 person-years in a 5-year follow-up study of Aboriginal Australians aged over 60 years,^46^ which is similar to our result (64.6 cases/1000 person-years). The observed sex- and age-disparities in incidence rates also corresponded to prior knowledge. For example, a review reported that women had a greater incidence rate of depression than men, and the incidence rate decreased after the 50s.^41^ Also, our results echoed the established knowledge that the risk of impairments in sensory, physical, and cognitive function was higher in older ages.^40 42 43 46 47 48^ Other than confirming previous findings, our study added valuable evidence of age- and sex-specific incidence rates of AFRIs in a wide range of age groups (51-90 years) within a cohort, which provided important evidence for the prevention of ARFIs in the aging era.

Based on these findings, we further looked into their patterns of co-existence and progression. Current literature has explored the co-existence of age-related diseases^16 17 18 49 50^ within the framework of multimorbidity. For example, a matched case-control study explored the co-existence and progression patterns of 5 combinations of diseases including pulmonary disease and metabolic disease in a claim database, which reported that aging was related to greater risk of and more complex patterns of comorbidity.^50^ We extended this framework to the ARFIs, which was also critical and directly related to the independence and well-being of older adults. Our findings implied that the aging process was a systematic deterioration of multiple functions, and healthy longevity requires holistic consideration of the intactness of multiple systems. Meanwhile, we discovered more complicated patterns of co-existence of ARFIs as age increased, which underscored the need for individualized early prevention, intervention, and care.

Taking a step forward, we constructed a hazard trajectory network of these ARFIs following an analysis framework of aging-related diseases.^15 16 17 18^ In 7.2 million Danish patients, Siggaard et al. constructed a disease trajectory browser to illustrate the connection network of diagnosed diseases.^16^ Using claim data, Jin and colleagues also conducted a network analysis for diseases with strong associations.^18^ We assessed the associations between ARFIs, based on previous studies that have discovered unidirectional^44 45 51 52^ or bidirectional relations between sensory impairments, such as vision and hearing, depression, and cognitive impairment.^44 45 51 53 54^ For example, a meta-analysis of longitudinal studies indicated that visual impairment was associated with a 66% higher risk of cognitive impairment, and that cognitive impairment was also associated with the incidence of visual impairment.^54^ As for risk of mortality, our findings also aligns with several long-term follow-up cohort studies.^20 21 23 24 25^ For instance, in a prospective analysis based on UK Biobank participants, physical frailty associated with 153% higher risk of all-cause mortality aged over 65 years.^20^ Based on these studies, we depicted a holistic view of the ARFIs and provided valuable evidence for the development of the comprehensive health management and targeted intervention.

Our findings have both clinical relevance and public health implications. Since ARFIs may be the result of multi-system pathophysiological changes,^55^ active and healthy aging should focus on the prevention of multiple function impairments as a whole.^56^ By preventing and delaying ARFIs, the goal of healthy longevity not merely prolongs life expectancy but also reduces disability among older adults.^57^ In clinical practice, diagnosis of a single function impairment may also imply other dysfunctions that are worth attention, thus helping in the early detection of other ARFIs and identification of individuals at higher mortality risk. Future research could combine functional changes, physiological mechanisms, and disease occurrence for early and precise identification of high-risk population.

Although the underlying mechanism remains unclear and complex, several possible pathways could connect changes from biological to phenotypic, and eventually functional aging.^58^ For example, studies have demonstrated that telomere shortening could indicate the cellular senescence,^59 60 61^ which was the basis for the initial biological aging. The accumulation of senescent cells may trigger functional changes in organs, and biological aging thereafter could cause phenotypic aging. For instance, the accumulation in the brain was related to the reduction in cognitive function,^62^ and the accumulation in bones and joints might disrupt the action function, increasing the risk of frailty and falls.^61^ Phenotypic aging may eventually lead to functional aging, e.g., macular degeneration related to telomerase activity was the most common cause of irreversible blindness in the elderly worldwide.^63^ Several studies have also provided evidence that telomeres were associated with changes in sensory function and neurocognitive function.^62 64 65^ Thus, the complex intertwining of ARFIs could be explained by certain specific mechanisms or pathways at different levels of aging.

Strengths of the current study included the prospective study design and long-term follow-up. The frequently measured functional phenotypes in the HRS over two decades provided a unique opportunity for our in-depth analyses, but our findings should be interpreted with caution due to some limitations. First, the ascertainment of ARFIs was based on self-reports or questionnaire^66^ and subject to measurement error, although previous studies showed the acceptable reproducibility and validity of the definitions.^27 28 30 31^ Second, we investigated six common ARFIs, while other AFRIs (e.g., olfactory impairment) were not included due to data limitations. Furthermore, since our focus was the functional impairments, we did not account for potential incident diseases such as cancer and stroke, which could be incorporated in future studies to more thoroughly reveal the aging process. Because our findings are based on a US cohort, their generalizability to other populations warrants confirmation. Finally, the hazard network does not necessarily indicate causal relations as residual confounding and reverse causation could still exist considering the observational nature of the current study.

## CONCLUSIONS

In this population-based study, the incidence rates of visual impairment, hearing impairment, physical frailty, and cognitive impairment increased exponentially as age increased, while incidence rates of restless sleep and depression increased relatively slowly with age. In a hazard trajectory network, all ARFIs were bidirectionally related to each other and predicted higher risk of mortality in a dose-response manner. Physical frailty, depression, visual impairment, and cognitive impairment independently predicted higher risk of mortality. Our study provides valuable evidence that ARFIs in older adults are highly correlated as a network, and that they are linked to higher risk of mortality. Therefore, integrated strategies for prevention and management of multiple ARFIs are needed to achieve healthy longevity.

## Data Availability

All data produced in the present study are available upon reasonable request to the authors.

## List of abbreviations

HRS: Health and Retirement Study
HR: hazard ratio
CI: confidence interval
SD: standard deviation
ARFI: age-related functional impairment
VI: visual impairment
HI: hearing impairment
CI: cognitive impairment
RS: restless sleep
PF: physical frailty

## Acknowledgments

The author would like to express genuine gratitude to the participants and staffs of the Health and Retirement Study, who contributed greatly to the academic community and made this study possible.

## Contributors

CY and HC designed the study; BW, HC, and TZ performed the statistical analyses; BW and TZ interpreted the data; HC and BW drafted the manuscript, RL, JS and XX further revised the manuscript; CY supervised the data analysis and interpretation; CY had the primary responsibility for the study final content. All authors critically reviewed the manuscript and approved the final draft.

## Ethical approval

This HRS was approved by the Institute for Social Research of University of Michigan (IRB Protocol: HUM00061128). Written informed consent was obtained from all participants or proxy.

## Data sharing

Data described in the manuscript and the codebook are available from the HRS website (https://hrs.isr.umich.edu/).

