## Supplemental materials for "Progression and trajectory network of age-related functional impairments and their associations with mortality: a two-decade prospective study"

### Supplementary tables and figures

**Table S1.** Age- and sex-specific incident rates of ARFIs, overall and by sex

**Table S2.** Co-existence pairs of age-related functional impairments in 2000, overall and by cohorts

**Table S3.** Prevalence of co-existence of ARFIs from 2000 to 2020, overall and by cohorts

**Table S4.** Hazard ratios for the associations between the ARFIs and mortality

**Table S5.** Sensitivity analysis of hazard trajectory network of ARFIs

**Figure S1.** Flow chart of the study population

**Figure S2.** Co-existence patterns of age-related functional impairments in 2000 by birth cohorts.

**Figure S3.** Hazard trajectory networks of age-related functional impairments

**Table S1. Age- and sex-specific incident rates of ARFIs, overall and by sex**

| **Age group** | **Incident rates (cases / 1000 person-years)** | | | | | |
| --- | --- | --- | --- | --- | --- | --- |
|  | **Visual impairment** | **Hearing impairment** | **Cognitive impairment** | **Physical frailty ^a^** | **Restless sleep** | **Depression** |
| **Both sexes** |  |  |  |  |  |  |
| 51-55 years | 46.8 | 29.8 | 23.0 | NA | 82.4 | 35.5 |
| 56-60 years | 42.6 | 34.1 | 29.3 | NA | 77.1 | 34.1 |
| 61-65 years | 50.3 | 43.5 | 39.4 | NA | 71.7 | 28.1 |
| 66-70 years | 57.7 | 53.8 | 45.0 | 20.7 | 69.1 | 34.5 |
| 71-75 years | 65.8 | 61.8 | 70.5 | 28.6 | 74.9 | 36.2 |
| 76-80 years | 85.7 | 82.4 | 91.5 | 38.6 | 79.4 | 40.1 |
| 81-85 years | 103.1 | 107.9 | 131.0 | 48.4 | 82.9 | 52.0 |
| 86-90 years | 145.4 | 131.5 | 187.5 | 62.9 | 87.3 | 74.9 |
| All ages | 59.3 | 52.1 | 42.5 | 31.7 | 75.6 | 35.6 |
| *P-trend* | < 0.001 | < 0.001 | < 0.001 | < 0.001 | < 0.001 | 0.781 |
| *P-nonlinear* | < 0.001 | 0.003 | 0.003 | 0.027 | < 0.001 | 0.044 |
| **Male** |  |  |  |  |  |  |
| 51-55 years | 42.6 | 42.8 | 24.9 | NA | 70.1 | 29.3 |
| 56-60 years | 39.8 | 47.3 | 30.6 | NA | 66.4 | 27.8 |
| 61-65 years | 48.9 | 62.0 | 42.5 | NA | 58.6 | 18.8 |
| 66-70 years | 56.1 | 69.6 | 44.2 | 13.2 | 56.8 | 25.1 |
| 71-75 years | 67.0 | 79.4 | 76.3 | 24.5 | 61.3 | 25.0 |
| 76-80 years | 84.4 | 100.0 | 94.1 | 27.8 | 71.5 | 31.4 |
| 81-85 years | 113.8 | 139.4 | 138.0 | 33.7 | 73.5 | 51.4 |
| 86-90 years | 140.6 | 149.1 | 162.9 | 55.3 | 88.2 | 82.8 |
| All ages | 57.2 | 67.4 | 41.6 | 22.8 | 63.6 | 27.3 |
| *P-trend* | < 0.001 | < 0.001 | < 0.001 | < 0.001 | < 0.001 | 0.519 |
| *P-nonlinear* | 0.071 | 0.354 | 0.206 | 0.327 | < 0.001 | 0.043 |
| **Female** |  |  |  |  |  |  |
| 51-55 years | 49.2 | 23.4 | 22.0 | NA | 89.4 | 38.8 |
| 56-60 years | 44.7 | 25.8 | 28.6 | NA | 84.8 | 38.4 |
| 61-65 years | 51.5 | 31.1 | 37.2 | NA | 82.3 | 35.4 |
| 66-70 years | 59.1 | 43.4 | 45.6 | 27.2 | 79.4 | 41.8 |
| 71-75 years | 64.9 | 51.3 | 66.4 | 32.0 | 85.6 | 44.9 |
| 76-80 years | 86.5 | 73.3 | 89.9 | 45.9 | 84.7 | 45.7 |
| 81-85 years | 97.7 | 95.2 | 127.2 | 57.0 | 88.5 | 52.3 |
| 86-90 years | 147.7 | 125.4 | 198.6 | 66.9 | 87.0 | 70.9 |
| All ages | 60.9 | 43.1 | 43.2 | 38.5 | 84.3 | 41.4 |
| *P-trend* | < 0.001 | < 0.001 | < 0.001 | < 0.001 | < 0.001 | 0.918 |
| *P-nonlinear* | <0.001 | 0.102 | 0.008 | 0.073 | 0.196 | 0.497 |

ARFI: age-related functional impairment, NA: not applicable.

Person-time was calculated from the visit in a specific age group to the next visit for each individual.

^a^ Participants aged below 65 were not measured for physical frailty.

**Table S2. Co-existence pairs of age-related functional impairments in 2000, overall and by cohorts**

|  | **VI** | **HI** | **CI** | **PF** | **RS** | **Depression** |
| --- | --- | --- | --- | --- | --- | --- |
| **All participants (N = 15 852, n (%))** | | | | | |  |
| **VI** | 3191 (20.1) |  |  |  |  |  |
| **HI** | 1116 (7.0) | 2925 (18.5) |  |  |  |  |
| **CI** | 921 (5.8) | 720 (4.5) | 2579 (16.3) |  |  |  |
| **PF** | 516 (3.3) | 391 (2.5) | 368 (2.3) | 1130 (7.1) |  |  |
| **RS** | 1469 (9.3) | 1179 (7.4) | 1021 (6.4) | 613 (3.9) | 5241 (33.1) |  |
| **Depression** | 781 (4.9) | 542 (3.4) | 568 (3.6) | 372 (2.3) | 1307 (8.2) | 1880 (11.9) |
| **WB cohort (N = 1829, n (%))** | | | | | | |
| **VI** | 290 (15.9) |  |  |  |  |  |
| **HI** | 64 (3.5) | 198 (10.8) |  |  |  |  |
| **CI** | 56 (3.1) | 23 (1.3) | 150 (8.2) |  |  |  |
| **RS** | 155 (8.5) | 80 (4.4) | 67 (3.7) | NA | 625 (34.2) |  |
| **Depression** | 68 (3.7) | 30 (1.6) | 45 (2.5) | NA | 153 (8.4) | 210 (11.5) |
| **HRS original cohort (N = 8365, n (%))** | | | | | |  |
| **VI** | 1504 (18.0) |  |  |  |  |  |
| **HI** | 475 (5.7) | 1326 (15.9) |  |  |  |  |
| **CI** | 348 (4.2) | 261 (3.1) | 960 (11.5) |  |  |  |
| **PF** | 151 (1.8) | 98 (1.2) | 78 (0.9) | 328 (3.9) |  |  |
| **RS** | 710 (8.5) | 556 (6.6) | 400 (4.8) | 190 (2.3) | 2739 (32.7) |  |
| **Depression** | 377 (4.5) | 244 (2.9) | 223 (2.7) | 114 (1.4) | 663 (7.9) | 929 (11.1) |
| **CODA cohort (N = 1892, n (%))** | | | | | |  |
| **VI** | 360 (19.0) |  |  |  |  |  |
| **HI** | 127 (6.7) | 370 (19.6) |  |  |  |  |
| **CI** | 135 (7.1) | 100 (5.3) | 386 (20.4) |  |  |  |
| **PF** | 82 (4.3) | 63 (3.3) | 62 (3.3) | 204 (10.8) |  |  |
| **RS** | 173 (9.1) | 158 (8.4) | 151 (8.0) | 115 (6.1) | 611 (32.3) |  |
| **Depression** | 84 (4.4) | 58 (3.1) | 88 (4.7) | 63 (3.3) | 149 (7.9) | 205 (10.8) |
| **AHEAD cohort (N = 3766, n (%))** | | | | | |  |
| **VI** | 1037 (27.5) |  |  |  |  |  |
| **HI** | 450 (11.9) | 1031 (27.4) |  |  |  |  |
| **CI** | 382 (10.1) | 336 (8.9) | 1083 (28.8) |  |  |  |
| **PF** | 283 (7.5) | 230 (6.1) | 228 (6.1) | 598 (15.9) |  |  |
| **RS** | 431 (11.4) | 385 (10.2) | 403 (10.7) | 308 (8.2) | 1266 (33.6) |  |
| **Depression** | 252 (6.7) | 210 (5.6) | 212 (5.6) | 195 (5.2) | 342 (9.1) | 536 (14.2) |

VI: visual impairment, HI: hearing impairment, CI: cognitive impairment, PF: physical frailty, RS: restless sleep, NA: not applicable, ARFI: age-related functional impairment, WB cohort: War Baby cohort born 1942 to 1947, HRS cohort: initial HRS cohort born 1931 to 1941, CODA cohort: Children of Depression cohort born 1924 to 1930, AHEAD cohort: Asset and Health Dynamics Among the Oldest Old cohort born before 1924.

PF was only measured among participants aged over 64 years.

**Table S3. Prevalence of co-existence of ARFIs from 2000 to 2020, overall and by cohorts**

| **Status** | **Follow-up Year** | | | | | | |
| --- | --- | --- | --- | --- | --- | --- | --- |
|  | **2000** | **2004** | **2008** | | **2012** | **2016** | **2020** |
| **All participants (N = 15 852, n (%))** | | | |  | | | |
| No ARFI | 6433 (40.6) | 4064 (25.6) | 2645 (16.7) | | 1846 (11.6) | 1196 (7.5) | 493 (3.1) |
| One ARFI | 4890 (30.8) | 3699 (23.3) | 2983 (18.8) | | 2230 (14.1) | 1628 (10.3) | 744 (4.7) |
| Two ARFIs | 2564 (16.2) | 2655 (16.7) | 2454 (15.5) | | 2014 (12.7) | 1501 (9.5) | 785 (5.0) |
| Three ARFIs | 1213 (7.7) | 1585 (10.0) | 1794 (11.3) | | 1627 (10.3) | 1220 (7.7) | 680 (4.3) |
| Four ARFIs | 516 (3.3) | 849 (5.4) | 849 (5.4) | | 832 (5.2) | 694 (4.4) | 424 (2.7) |
| Five ARFIs | 191 (1.2) | 355 (2.2) | 395 (2.5) | | 390 (2.5) | 297 (1.9) | 235 (1.5) |
| Six ARFIs | 45 (0.3) | 84 (0.5) | 138 (0.9) | | 135 (0.9) | 104 (0.7) | 97 (0.6) |
| Not visited | 0 (0.0) | 1132 (7.1) | 1317 (8.3) | | 1494 (9.4) | 2085 (13.2) | 4450 (28.1) |
| Death | 0 (0.0) | 1429 (9.0) | 3277 (20.7) | | 5284 (33.3) | 7127 (45.0) | 7944 (50.1) |
| **WB cohort (N = 1829, n (%))** | | | |  | | | |
| No ARFI | 886 (48.4) | 672 (36.7) | 528 (28.9) | | 406 (22.2) | 308 (16.8) | 138 (7.5) |
| One ARFI | 576 (31.5) | 482 (26.4) | 446 (24.4) | | 391 (21.4) | 330 (18.0) | 166 (9.1) |
| Two ARFIs | 244 (13.3) | 289 (15.8) | 299 (16.3) | | 303 (16.6) | 264 (14.4) | 148 (8.1) |
| Three ARFIs | 91 (5.0) | 127 (6.9) | 160 (8.7) | | 189 (10.3) | 167 (9.1) | 117 (6.4) |
| Four ARFIs | 24 (1.3) | 55 (3.0) | 76 (4.2) | | 81 (4.4) | 82 (4.5) | 63 (3.4) |
| Five ARFIs | 8 (0.4) | 22 (1.2) | 32 (1.7) | | 31 (1.7) | 40 (2.2) | 27 (1.5) |
| Six ARFIs | 0 (0.0) | 0 (0.0) | 2 (0.1) | | 20 (1.1) | 12 (0.7) | 19 (1.0) |
| Not visited | 0 (0.0) | 142 (7.8) | 184 (10.1) | | 231 (12.6) | 346 (18.9) | 817 (44.7) |
| Death | 0 (0.0) | 40 (2.2) | 102 (5.6) | | 177 (9.7) | 280 (15.3) | 334 (18.3) |
| **HRS original cohort (N = 8365, n (%))** | | | |  | | | |
| No ARFI | 3784 (45.2) | 2617 (31.3) | 1769 (21.1) | | 1281 (15.3) | 821 (9.8) | 345 (4.1) |
| One ARFI | 2572 (30.7) | 2069 (24.7) | 1841 (22.0) | | 1474 (17.6) | 1137 (13.6) | 533 (6.4) |
| Two ARFIs | 1197 (14.3) | 1344 (16.1) | 1359 (16.2) | | 1232 (14.7) | 987 (11.8) | 556 (6.6) |
| Three ARFIs | 518 (6.2) | 787 (9.4) | 980 (11.7) | | 948 (11.3) | 801 (9.6) | 465 (5.6) |
| Four ARFIs | 217 (2.6) | 368 (4.4) | 428 (5.1) | | 462 (5.5) | 428 (5.1) | 294 (3.5) |
| Five ARFIs | 64 (0.8) | 159 (1.9) | 190 (2.3) | | 232 (2.8) | 189 (2.3) | 172 (2.1) |
| Six ARFIs | 13 (0.2) | 37 (0.4) | 76 (0.9) | | 77 (0.9) | 72 (0.9) | 58 (0.7) |
| Not visited | 0 (0.0) | 557 (6.7) | 702 (8.4) | | 845 (10.1) | 1246 (14.9) | 2812 (33.6) |
| Death | 0 (0.0) | 427 (5.1) | 1020 (12.2) | | 1814 (21.7) | 2684 (32.1) | 3130 (37.4) |
| **CODA cohort (N = 1892, n (%))** | | | |  | | | |
| No ARFI | 740 (39.1) | 404 (21.4) | 191 (10.1) | | 113 (6.0) | 51 (2.7) | 7 (0.4) |
| One ARFI | 581 (30.7) | 458 (24.2) | 348 (18.4) | | 221 (11.7) | 106 (5.6) | 36 (1.9) |
| Two ARFIs | 311 (16.4) | 314 (16.6) | 304 (16.1) | | 222 (11.7) | 154 (8.1) | 49 (2.6) |
| Three ARFIs | 154 (8.1) | 195 (10.3) | 229 (12.1) | | 211 (11.2) | 124 (6.6) | 65 (3.4) |
| Four ARFIs | 70 (3.7) | 108 (5.7) | 109 (5.8) | | 127 (6.7) | 93 (4.9) | 38 (2.0) |
| Five ARFIs | 25 (1.3) | 53 (2.8) | 60 (3.2) | | 56 (3.0) | 36 (1.9) | 24 (1.3) |
| Six ARFIs | 11 (0.6) | 12 (0.6) | 22 (1.2) | | 15 (0.8) | 9 (0.5) | 12 (0.6) |
| Not visited | 0 (0.0) | 145 (7.7) | 160 (8.5) | | 167 (8.8) | 235 (12.4) | 426 (22.5) |
| Death | 0 (0.0) | 203 (10.7) | 469 (24.8) | | 760 (40.2) | 1084 (57.3) | 1235 (65.3) |
| **AHEAD cohort (N = 3766, n (%))** | | | |  | | | |
| No ARFI | 1023 (27.2) | 371 (9.9) | 157 (4.2) | | 46 (1.2) | 16 (0.4) | 3 (0.1) |
| One ARFI | 1161 (30.8) | 690 (18.3) | 348 (9.2) | | 144 (3.8) | 55 (1.5) | 9 (0.2) |
| Two ARFIs | 812 (21.6) | 708 (18.8) | 492 (13.1) | | 257 (6.8) | 96 (2.5) | 32 (0.8) |
| Three ARFIs | 450 (11.9) | 476 (12.6) | 425 (11.3) | | 279 (7.4) | 128 (3.4) | 33 (0.9) |
| Four ARFIs | 205 (5.4) | 318 (8.4) | 236 (6.3) | | 162 (4.3) | 91 (2.4) | 29 (0.8) |
| Five ARFIs | 94 (2.5) | 121 (3.2) | 113 (3.0) | | 71 (1.9) | 32 (0.8) | 12 (0.3) |
| Six ARFIs | 21 (0.6) | 35 (0.9) | 38 (1.0) | | 23 (0.6) | 11 (0.3) | 8 (0.2) |
| Not visited | 0 (0.0) | 288 (7.6) | 271 (7.2) | | 251 (6.7) | 258 (6.9) | 395 (10.5) |
| Death | 0 (0.0) | 759 (20.2) | 1686 (44.8) | | 2533 (67.3) | 3079 (81.8) | 3245 (86.2) |

ARFI: age-related functional impairment, WB cohort: War Baby cohort born 1942 to 1947, HRS cohort: initial HRS cohort born 1931 to 1941, CODA cohort: Children of Depression cohort born 1924 to 1930, AHEAD cohort: Asset and Health Dynamics Among the Oldest Old cohort born before 1924.

**Table S4. Hazard ratios for the associations between the ARFIs and mortality**

| Exposure | Outcome | Cases/N | HR (95% CI) | P-value |
| --- | --- | --- | --- | --- |
| Visual impairment | Hearing impairment | 2043/4781 | 2.48 [2.29, 2.69] | < 0.001 |
|  | Cognitive impairment | 3117/5704 | 1.63 [1.53, 1.74] | < 0.001 |
|  | Depression | 1696/5458 | 2.18 [2.00, 2.39] | < 0.001 |
|  | Physical frailty | 2518/6080 | 2.08 [1.92, 2.24] | < 0.001 |
|  | Restless sleep | 1717/3559 | 1.72 [1.59, 1.86] | < 0.001 |
|  | Mortality | 5287/9463 | 1.19 [1.13, 1.26] | < 0.001 |
| Hearing impairment | Visual impairment | 1980/4044 | 2.10 [1.95, 2.27] | < 0.001 |
|  | Cognitive impairment | 2858/5323 | 1.37 [1.29, 1.46] | < 0.001 |
|  | Depression | 1429/5202 | 1.56 [1.42, 1.70] | < 0.001 |
|  | Physical frailty | 2252/5773 | 1.67 [1.55, 1.81] | < 0.001 |
|  | Restless sleep | 1583/3455 | 1.43 [1.32, 1.55] | < 0.001 |
|  | Mortality | 4818/8610 | 0.95 [0.90, 1.00] | 0.073 |
| Cognitive impairment | Visual impairment | 1894/4006 | 1.72 [1.57, 1.88] | < 0.001 |
|  | Hearing impairment | 1705/4427 | 1.43 [1.30, 1.57] | < 0.001 |
|  | Depression | 1262/4790 | 1.50 [1.35, 1.68] | < 0.001 |
|  | Physical frailty | 2139/5485 | 1.56 [1.42, 1.71] | < 0.001 |
|  | Restless sleep | 1268/3202 | 1.25 [1.13, 1.38] | < 0.001 |
|  | Mortality | 5019/8678 | 1.13 [1.06, 1.21] | < 0.001 |
| Depression | Visual impairment | 1290/2445 | 2.04 [1.89, 2.21] | < 0.001 |
|  | Hearing impairment | 1159/2952 | 1.71 [1.57, 1.86] | < 0.001 |
|  | Cognitive impairment | 1860/3500 | 1.51 [1.41, 1.61] | < 0.001 |
|  | Physical frailty | 1569/3205 | 2.31 [2.14, 2.49] | < 0.001 |
|  | Restless sleep | 622/1097 | 2.41 [2.17, 2.67] | < 0.001 |
|  | Mortality | 2844/5341 | 1.38 [1.30, 1.46] | < 0.001 |
| Physical frailty | Visual impairment | 1277/2651 | 2.08 [1.90, 2.29] | < 0.001 |
|  | Hearing impairment | 1153/3170 | 1.87 [1.70, 2.06] | < 0.001 |
|  | Cognitive impairment | 1949/3756 | 1.62 [1.50, 1.75] | < 0.001 |
|  | Depression | 1008/3040 | 2.95 [2.65, 3.28] | < 0.001 |
|  | Restless sleep | 784/1625 | 2.14 [1.92, 2.38] | < 0.001 |
|  | Mortality | 4078/6780 | 1.59 [1.49, 1.69] | < 0.001 |
| Restless sleep | Visual impairment | 2874/6365 | 1.43 [1.34, 1.53] | < 0.001 |
|  | Hearing impairment | 2619/3838 | 1.43 [1.33, 1.54] | < 0.001 |
|  | Cognitive impairment | 3866/7963 | 1.12 [1.06, 1.19] | < 0.001 |
|  | Depression | 2318/7024 | 3.67 [3.32, 4.05] | < 0.001 |
|  | Physical frailty | 3228/7787 | 1.74 [1.61, 1.87] | < 0.001 |
|  | Mortality | 5038/10 444 | 1.05 [1.00, 1.11] | 0.052 |

All analyses were adjusted for age, gender, BMI, race, education, family income, smoking status, drinking status and vigorous exercise at baseline and other baseline and incident ARFIs.

**Table S5. Sensitivity analysis of hazard trajectory network of ARFIs**

| Exposure | Outcome | Sensitivity analysis 1^a^ | | Sensitivity analysis 2^b^ | | Sensitivity analysis 3^c^ | |
| --- | --- | --- | --- | --- | --- | --- | --- |
|  |  | HR (95% CI) | P-value | HR (95% CI) | P-value | HR (95% CI) | P-value |
| Visual impairment | Hearing impairment | 2.49 [2.27, 2.72] | < 0.001 | 2.06 [1.70, 2.50] | < 0.001 | 2.23 [2.06, 2.41] | < 0.001 |
|  | Cognitive impairment | 1.61 [1.50, 1.74] | < 0.001 | 1.58 [1.35, 1.84] | < 0.001 | 1.43 [1.34, 1.52] | < 0.001 |
|  | Depression | 2.18 [1.96, 2.42] | < 0.001 | 2.40 [1.91, 3.02] | < 0.001 | 1.97 [1.81, 2.15] | < 0.001 |
|  | Physical frailty | 1.99 [1.82, 2.18] | < 0.001 | 1.83 [1.53, 2.19] | < 0.001 | 1.94 [1.80, 2.09] | < 0.001 |
|  | Restless sleep | 1.61 [1.47, 1.77] | < 0.001 | 1.71 [1.42, 2.07] | < 0.001 | 1.59 [1.47, 1.71] | < 0.001 |
|  | Mortality | 1.14 [1.06, 1.22] | < 0.001 | 1.04 [0.90, 1.21] | 0.590 |  | < 0.001 |
| Hearing impairment | Visual impairment | 2.12 [1.94, 2.31] | < 0.001 | 1.81 [1.50, 2.19] | < 0.001 | 2.02 [1.87, 2.17] | < 0.001 |
|  | Cognitive impairment | 1.40 [1.30, 1.50] | < 0.001 | 1.40 [1.20, 1.63] | < 0.001 | 1.32 [1.24, 1.40] | < 0.001 |
|  | Depression | 1.58 [1.42, 1.75] | < 0.001 | 1.56 [1.23, 1.97] | < 0.001 | 1.44 [1.32, 1.57] | < 0.001 |
|  | Physical frailty | 1.74 [1.59, 1.90] | < 0.001 | 1.51 [1.25, 1.81] | < 0.001 | 1.69 [1.57, 1.82] | < 0.001 |
|  | Restless sleep | 1.43 [1.31, 1.57] | < 0.001 | 1.46 [1.21, 1.76] | < 0.001 | 1.40 [1.30, 1.50] | < 0.001 |
| Cognitive impairment | Visual impairment | 1.78 [1.60, 1.97] | < 0.001 | 1.53 [1.26, 1.86] | < 0.001 | 1.56 [1.43, 1.70] | < 0.001 |
|  | Hearing impairment | 1.45 [1.30, 1.62] | < 0.001 | 1.53 [1.24, 1.89] | < 0.001 | 1.27 [1.16, 1.40] | < 0.001 |
|  | Depression | 1.53 [1.34, 1.74] | < 0.001 | 1.29 [0.97, 1.70] | 0.076 | 1.37 [1.23, 1.52] | < 0.001 |
|  | Physical frailty | 1.61 [1.45, 1.80] | < 0.001 | 1.47 [1.20, 1.81] | < 0.001 | 1.46 [1.34, 1.60] | < 0.001 |
|  | Restless sleep | 1.23 [1.10, 1.38] | < 0.001 | 1.21 [0.97, 1.50] | 0.086 | 1.18 [1.07, 1.29] | < 0.001 |
|  | Mortality | 1.17 [1.08, 1.27] | < 0.001 | 1.07 [0.92, 1.25] | 0.354 |  | < 0.001 |
| Depression | Visual impairment | 1.89 [1.72, 2.08] | < 0.001 | 1.90 [1.52, 2.39] | < 0.001 | 1.76 [1.63, 1.9] | < 0.001 |
|  | Hearing impairment | 1.60 [1.44, 1.78] | < 0.001 | 1.47 [1.15, 1.89] | 0.002 | 1.45 [1.33, 1.58] | < 0.001 |
|  | Cognitive impairment | 1.46 [1.34, 1.59] | < 0.001 | 1.80 [1.49, 2.17] | < 0.001 | 1.20 [1.12, 1.29] | < 0.001 |
|  | Physical frailty | 2.13 [1.94, 2.35] | < 0.001 | 2.07 [1.67, 2.57] | < 0.001 | 2.03 [1.87, 2.19] | < 0.001 |
|  | Restless sleep | 2.08 [1.83, 2.37] | < 0.001 | 1.93 [1.42, 2.63] | < 0.001 | 2.06 [1.86, 2.28] | < 0.001 |
|  | Mortality | 1.31 [1.22, 1.42] | < 0.001 | 1.40 [1.15, 1.69] | < 0.001 |  | < 0.001 |
| Physical frailty | Visual impairment | 1.98 [1.76, 2.22] | < 0.001 | 1.91 [1.47, 2.49] | < 0.001 | 1.77 [1.61, 1.94] | < 0.001 |
|  | Hearing impairment | 1.83 [1.62, 2.06] | < 0.001 | 2.45 [1.85, 3.23] | < 0.001 | 1.54 [1.40, 1.70] | < 0.001 |
|  | Cognitive impairment | 1.50 [1.36, 1.65] | < 0.001 | 1.61 [1.30, 2.00] | < 0.001 | 1.28 [1.18, 1.38] | < 0.001 |
|  | Depression | 2.83 [2.48, 3.24] | < 0.001 | 2.94 [2.10, 4.10] | < 0.001 | 2.36 [2.12, 2.63] | < 0.001 |
|  | Restless sleep | 2.00 [1.75, 2.29] | < 0.001 | 1.61 [1.17, 2.22] | 0.004 | 1.81 [1.63, 2.01] | < 0.001 |
|  | Mortality | 1.59 [1.47, 1.73] | < 0.001 | 1.70 [1.40, 2.05] | < 0.001 |  | < 0.001 |
| Restless sleep | Visual impairment | 1.40 [1.30, 1.52] | < 0.001 | 1.31 [1.12, 1.54] | 0.001 | 1.40 [1.31, 1.49] | < 0.001 |
|  | Hearing impairment | 1.42 [1.30, 1.54] | < 0.001 | 1.16 [0.98, 1.38] | 0.086 | 1.39 [1.30, 1.49] | < 0.001 |
|  | Cognitive impairment | 1.12 [1.05, 1.20] | < 0.001 | 1.20 [1.04, 1.37] | 0.010 | 1.08 [1.02, 1.14] | 0.011 |
|  | Depression | 3.58 [3.19, 4.02] | < 0.001 | 2.95 [2.40, 3.62] | < 0.001 | 3.56 [3.23, 3.93] | < 0.001 |
|  | Physical frailty | 1.73 [1.59, 1.88] | < 0.001 | 1.75 [1.50, 2.05] | < 0.001 | 1.75 [1.63, 1.88] | < 0.001 |

All analyses were adjusted for age, gender, BMI, race, education, family income, smoking status, drinking status and vigorous exercise at baseline and other baseline and incident ARFIs.

^a^ In sensitivity analysis 1, we excluded participants with prevalent stroke, cancer, memory-related diseases or other psychological diseases

^b^ In sensitivity analysis 2, we restricted analyses in participants with no ARFI at baseline.

^c^ In sensitivity analysis 3, we used Fine and Grey’s competing risk models in place of Cox models, treating death as the competing risk.

**Figure S1. Flow chart of the study population**

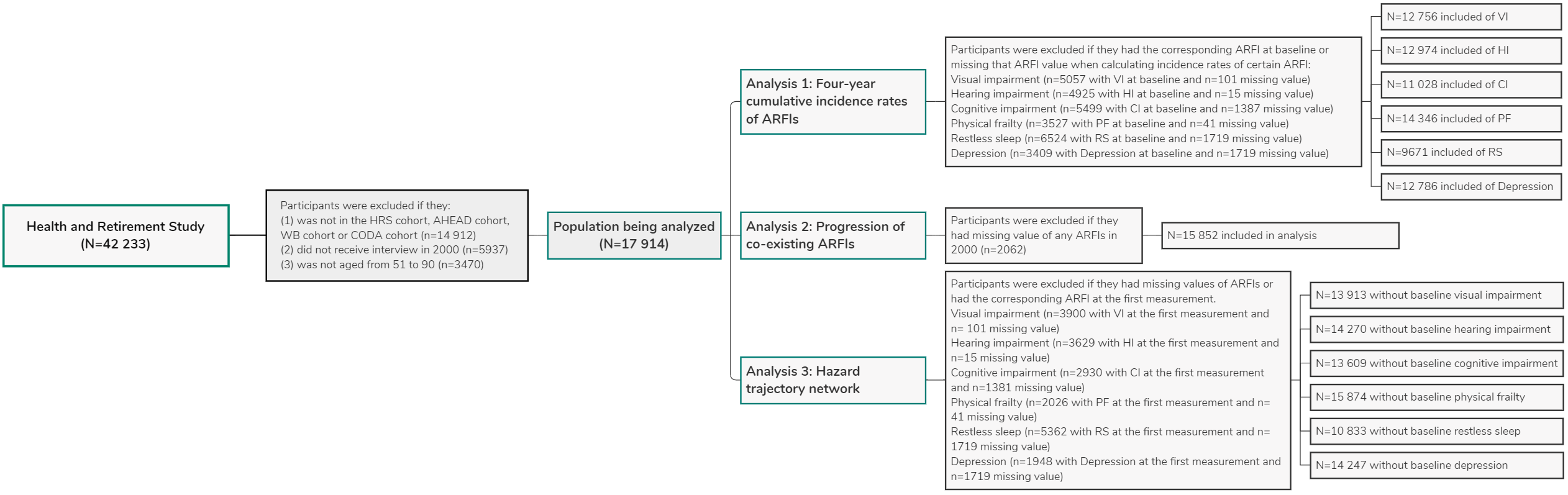

ARFI: age-related functional impairment, VI: visual impairment, HI: hearing impairment, CI: cognitive impairment, PF: physical frailty, RS: restless sleep, WB: War Baby cohort born in 1942 to 1947, HRS: initial HRS cohort born in 1931 to 1941, CODA: Children of Depression cohort born in 1924 to 1930, AHEAD: Asset and Health Dynamics Among the Oldest Old cohort born before 1924.

**Figure S2. Co-existence patterns of age-related functional impairments in 2000 by birth cohorts.**

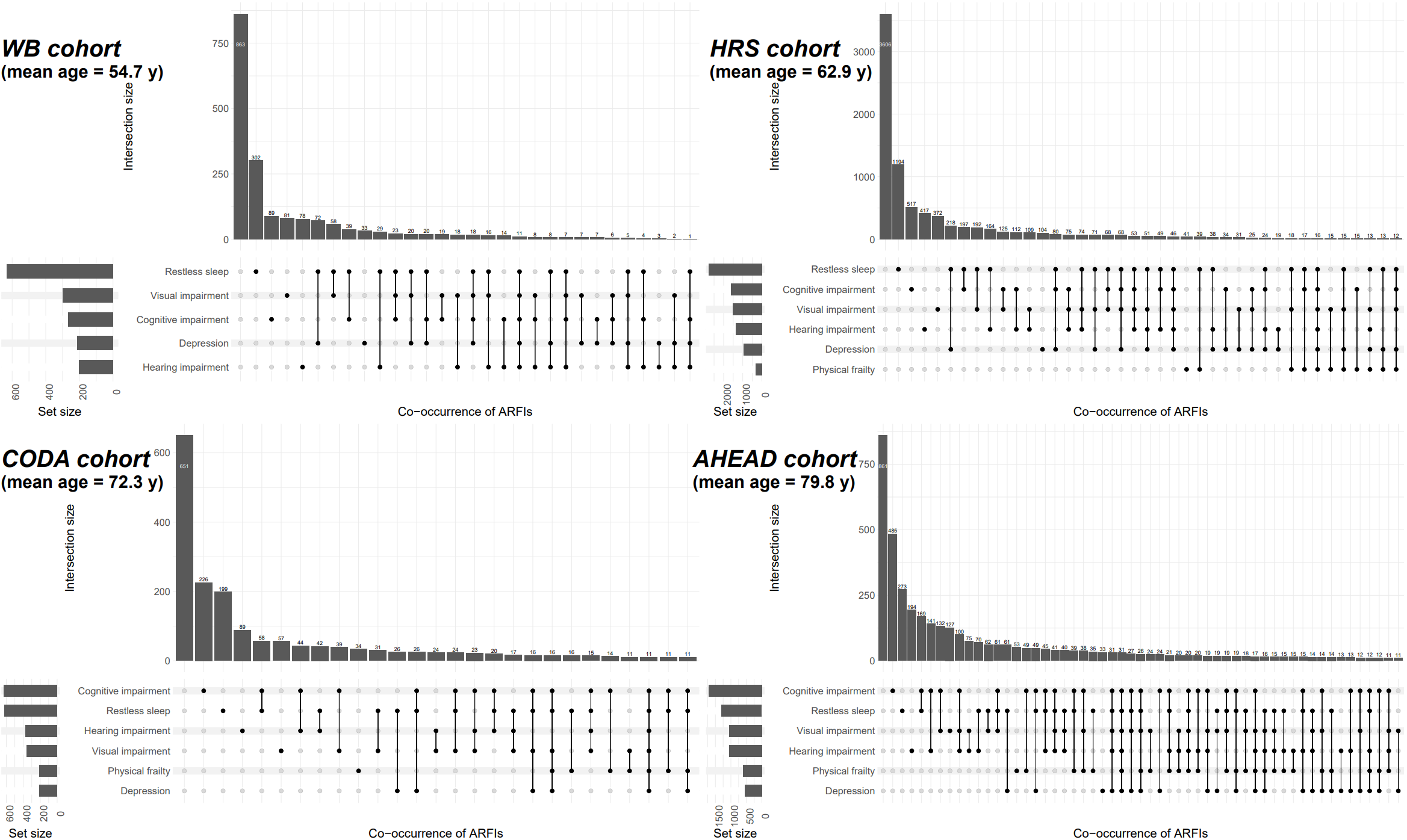

WB cohort: War Baby cohort born 1942 to 1947; HRS cohort: HRS original cohort born 1931 to 1941; CODA cohort: Children of Depression cohort born 1924 to 1930; AHEAD cohort: Asset and Health Dynamics Among the Oldest Old cohort born before 1924. The analysis included 1,657 participants from the WB cohort, 8,427 from HRS original cohort, 1,850 from the CODA cohort, and 3,768 from the AHEAD cohort. Combinations with <=10 cases were omitted.

**Figure S3. Hazard trajectory networks of age-related functional impairments**

A. **Initial analysis**

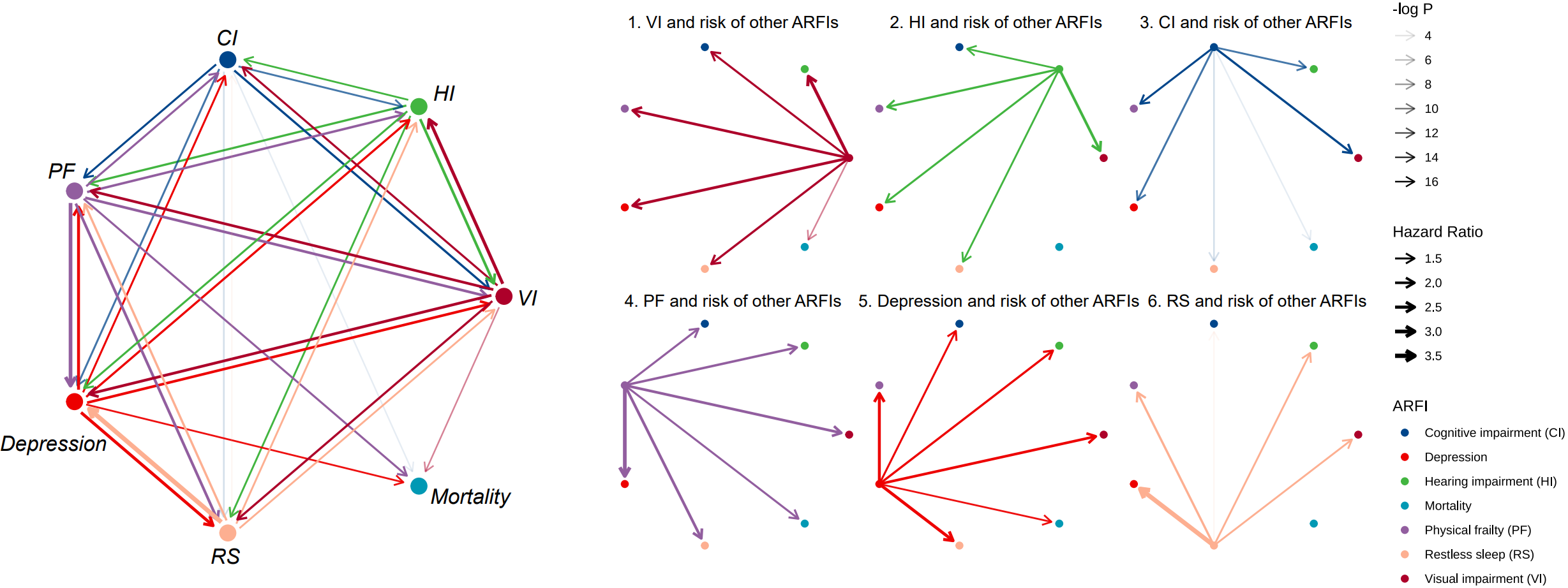

B. **Sensitivity analysis 1**

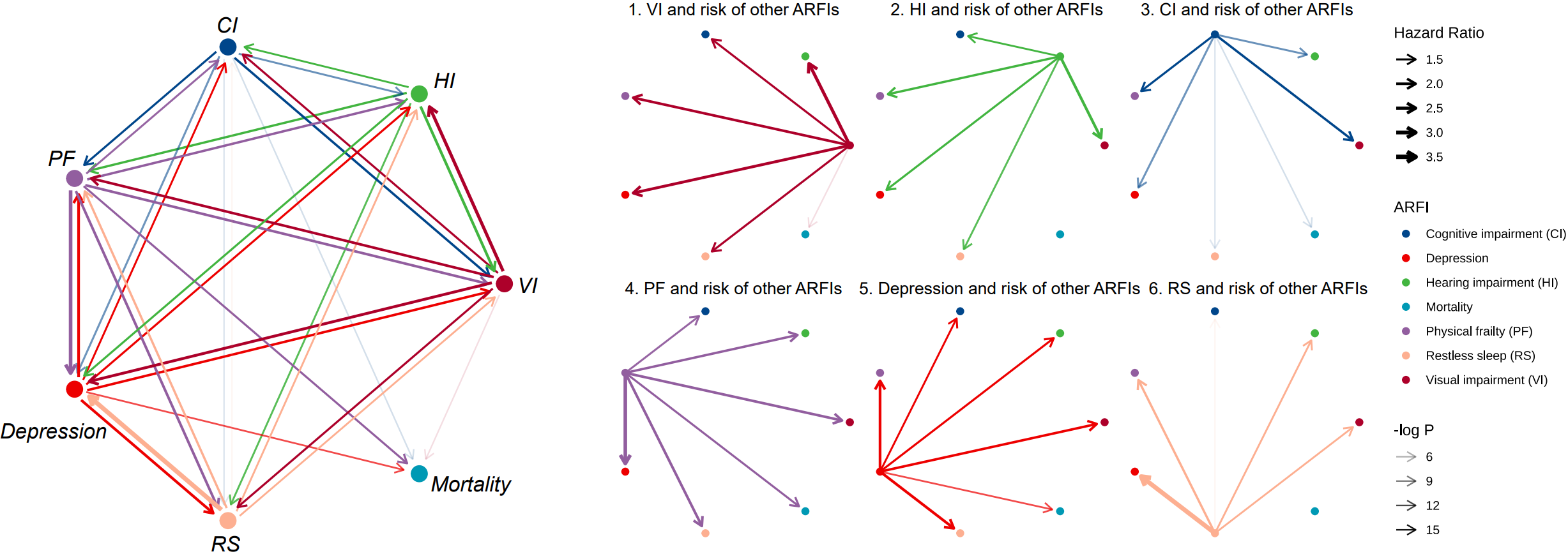

C. **Sensitivity analysis 2**

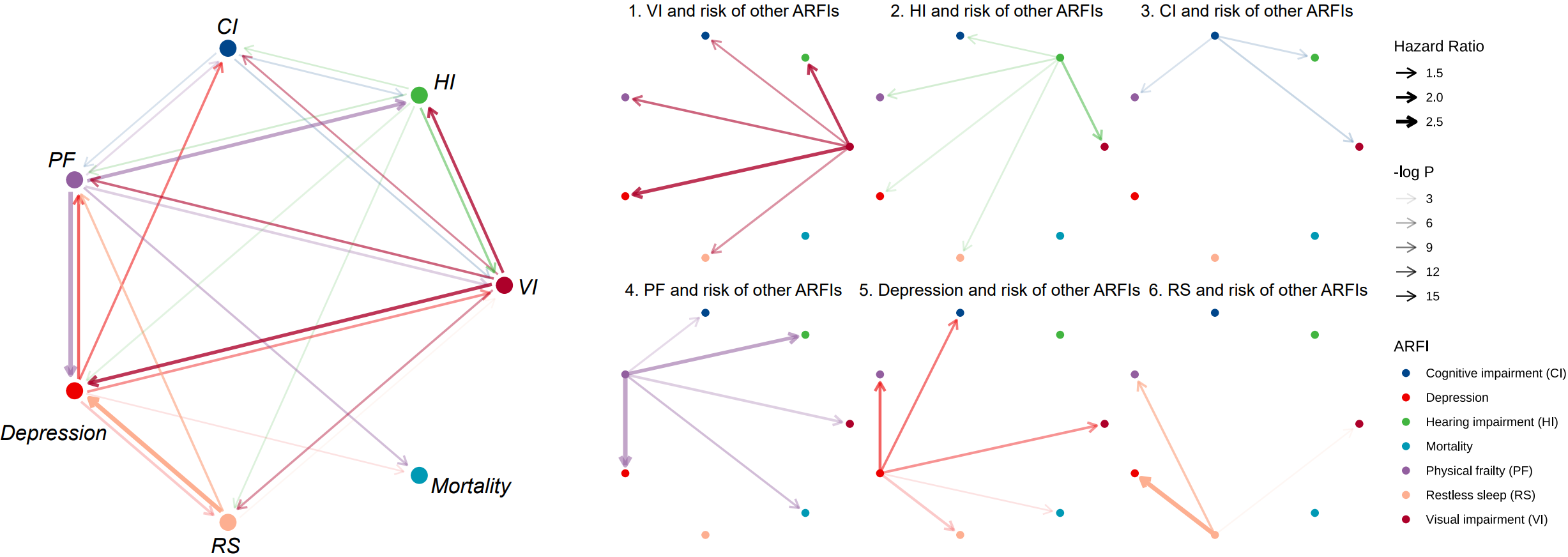

D. **Sensitivity analysis 3**

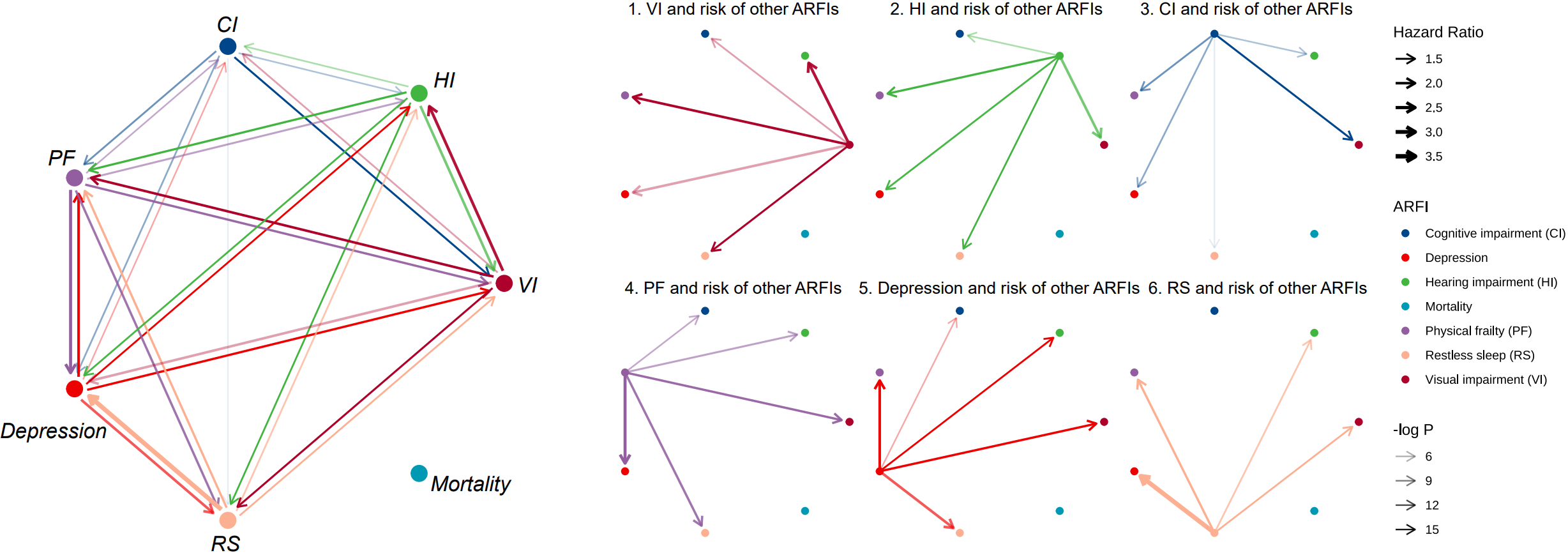

ARFI: age-related functional impairment, VI: visual impairment, HI: hearing impairment, CI: cognitive impairment, PF: physical frailty, RS: restless sleep.

All analyses were adjusted for age, gender, BMI, race, education, family income, smoking status, drinking status and vigorous exercise at baseline and other baseline and incident ARFIs.

In sensitivity analysis 1, we excluded participants with prevalent stroke, cancer, memory-related diseases, or other psychological diseases

In sensitivity analysis 2, we restricted analyses in participants with no ARFI at baseline.

In sensitivity analysis 3, we used Fine and Grey’s competing risk models in place of Cox models, treating death as the competing risk.
